# Electrostatic filters to reduce COVID-19 spread in bubble CPAP: an *in vitro* study of safety and efficacy

**DOI:** 10.1101/2020.09.23.20200485

**Authors:** Jonathan W Davis, J Jane Pillow, Matt N Cooper, Mar Janna Dahl

## Abstract

**Background:** Bubble CPAP may be used in infants with suspected or confirmed COVID-19. Electrostatic filters may reduce cross-infection. This study aims to determine if including a filter in the bubble CPAP circuit impacts stability of pressure delivery.

**Methods:** A new electrostatic filter was placed before (pre) or after (post) the bubble CPAP generator, or with no filter (control) in an *in vitro* study. Pressure was recorded at the nasal interface for 18 h (6 L/min; 7 cmH_2_O) on three occasions for each configuration. Filter failure was defined as pressure >9 cmH_2_0 for 60 continuous minutes. The filter was weighed before and after each experiment.

**Results:** Mean (SD) time to reach the fail-point was 257 (116) min and 525 (566) min for filter placement pre- and post-CPAP generator, respectively. Mean pressure was higher throughout in the pre-generator position compared to control. The filter weight was heavier at study end in the pre-compared to the post-generator position.

**Conclusions:** Placement of the filter at the pre-generator position in a bubble CPAP circuit should be avoided due to unstable mean pressure. Filters are likely to become saturated with water over time. The post-generator position may accommodate a filter, but regular pressure monitoring and early replacement are required.

## Introduction

SARS-CoV-2 originated in Wuhan, China, in November 2020 [1]. The clinical disease, Covid-19 was declared a global pandemic by WHO in March 2020 [2]. Data on neonatal infection with SARS-CoV-2 are limited [3-5]. The need for respiratory support in infants with Covid-19 is uncertain.

Continuous positive airway pressure (CPAP) has been a cornerstone treatment of neonatal respiratory distress syndrome for more than 40 years. CPAP use has extended in recent years to include infants with viral respiratory disease [6]. Bubble CPAP is a popular means of providing respiratory support in these infants. The expiratory limb of the bubble CPAP circuit vents through an underwater seal [7], creating bubbles. The resulting pressure oscillations are transmitted back to the nares and to the lung, hence the bubbling delivers a variable rather than constant pressure [8]. These pressure oscillations (frequency) may improve acute respiratory mechanics compared to constant delivery pressure consequent to recruitment of atelectatic alveoli [7-11].

CPAP generates aerosols and therefore, may promote transmission of viral particles [12, 13]. Electrostatic filters remove viruses from expired gas, offering protection to health-care staff. Manufacturers of electrostatic filters recommend electrostatic filter replacement every 24 h [14]. We hypothesised that an electrostatic filter placed in the humidified bias flow would become moisture saturated, resulting in instability in pressure transmission during non-invasive respiratory support. We aimed to determine how the placement and position of an electrostatic filter in a bubble CPAP circuit impacts the stability of the delivered pressure in an *in-vitro* model.

## Methods

An *in vitro* study was conducted at the University of Western Australia in April 2020. A simulated *in vitro* neonatal lung on a bubble CPAP model was created by connecting a test lung (Dräger, Lubeck, Germany) to a bubble CPAP generator (Fisher & Paykel, Auckland, New Zealand). An infant nasal CPAP cannula (Size 4, Hudson RCI, Teleflex, NC USA;) was interfaced between the bias flow circuit and the test lung. Humidified bias flow (MR290; Fisher & Paykel, Auckland, New Zealand) was set at 6 L/min (of air). Continuous positive airway pressure was measured at the proximal pressure line connector of the CPAP cannula using a physiological pressure transducer and dome (MLT844, AD Instruments, Australia). The bubble CPAP generator and humidifier were set below the test lung, which was maintained on a radiant-heated neonatal cot (Cosy-Cot; Fisher and Paykel, Auckland, New Zealand). The cot warmer was set at 75 %. Environmental ambient temperature was 23.9-24.7 °C throughout the recording periods.

### Filter Placement

A DAR(tm) pediatric-neonatal electrostatic filter (size small; Covidien, Mansfield, MA USA) was placed at one of two points in the breathing circuit: on the CPAP probe immediately before the exit of the bias flow into the water column (pre-generator; Figure 1B) or at the bias flow exit point, normally used as the water inlet (post-generator; Figure 1C). A new filter was used for each experimental repeat and was weighed before and after each trial. A bubble CPAP circuit without a filter was used as the control (Figure 1A) for the two experimental arms. Each filter position and control were repeated three times.

**Figure 1.**
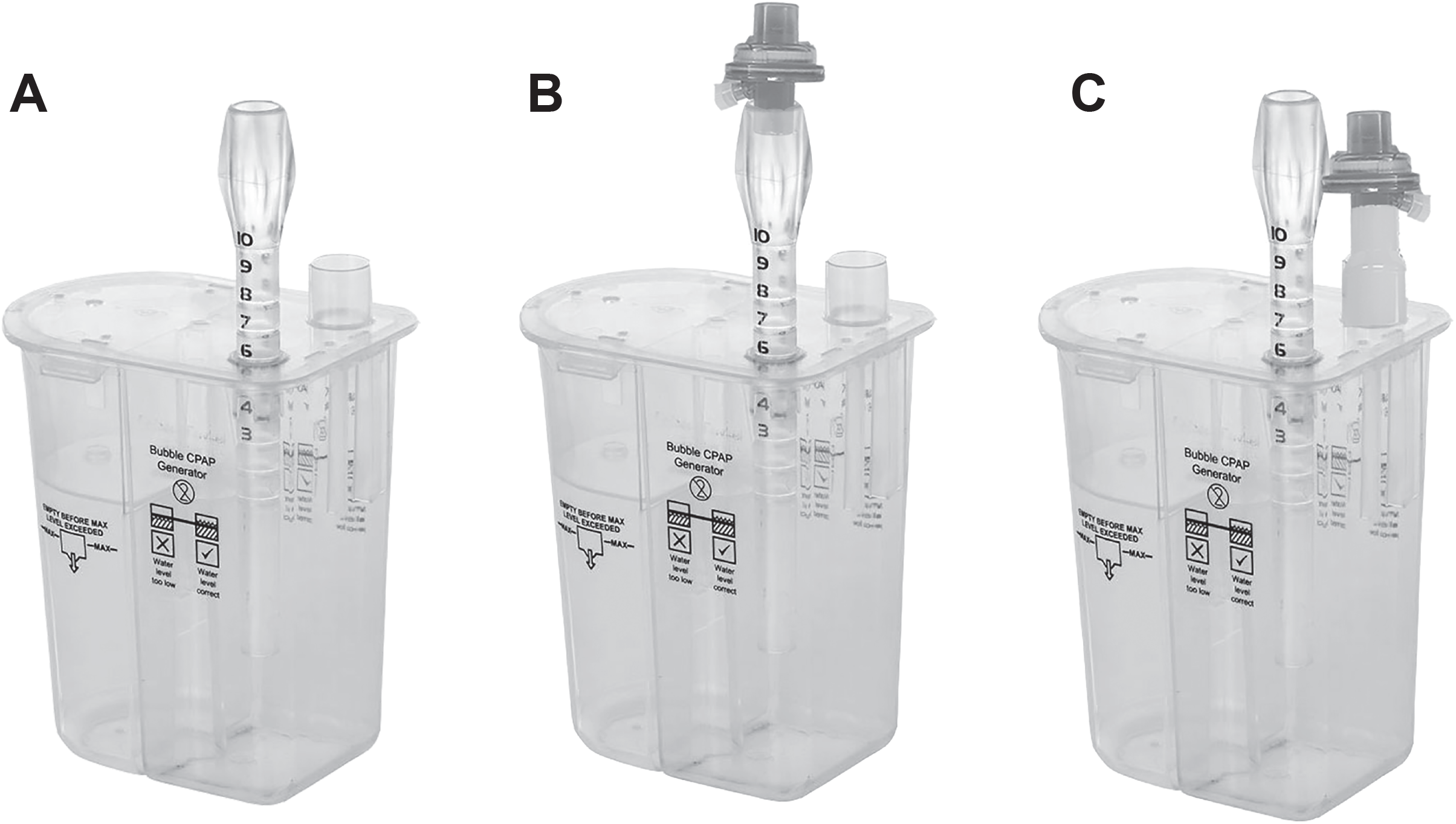
Filter placement. A) Control, standard set up; B) pre-generator expiratory limb; and C) post-generator at point of flow exit into environment.

### Signal Recording

The pressure signal was amplified, digitised (ML880; PowerLab ADInstruments, Bella Vista, NSW, Australia), and recorded continuously for 18 hours (LabChart 8, ADInstruments, Bella Vista, NSW, Australia) onto a laptop computer. Pressure and frequency data were collected at 5-minute intervals in 10-second epochs at a sampling frequency of 1 kHz.

### Analysis

The filter position fail-point was defined as a mean pressure exceeding 9 cmH_2_O for more than 60 continuous minutes. Mean pressure variability in a no-leak bubble CPAP system is 25 % greater than the set pressure [15]. Our fail-point allowed a 25 % increase on top of the set pressure to the nearest integer. The mean pressure and pressure difference (ΔP) of the bubble oscillations for each group (pre-generator, post-generator, and control) were compared across the 18 hours using a two-way repeated-measures ANOVA with Holm-Sidak *posthoc* correction. The frequency oscillations, unique to bubble CPAP, were assessed by transforming the mean pressure time series recording into the frequency domain using power spectral analysis. The mean frequency at the maximum power signal of the three circuit configurations was compared using a two-way repeated-measures ANOVA with Holm-Sidak *posthoc* correction. Filter weight and water volume in the circuit and overflow were compared by an unpaired Student t-test and one-way ANOVA, respectively. P-values were assessed at a 0.05 significance level by SigmaPlot (Systat, Inc., Santa Clara, CA)

## Results

### CPAP Fail-point

Failure to maintain mean pressure below 9 cmH_2_O occurred for 3 tests of the pre-generator and 2 of the 3 tests of the post-generator position. The mean (SD) time to the fail-point in pre-generator position was 257 (116) minutes and 525 (566) minutes for post-generator position. Mean pressure did not reach the fail-point in the control group.

### Mean pressure, ΔP, and frequency

The mean pressure with the filter in the pre-generator position increased progressively throughout the experiment and was significantly greater than control in 15 of the 18 hours of the study. ΔP was lower with the filter in the pre-generator position from three hours after study commencement. In contrast, the increase in mean pressure with the filter at the post-generator position was not significant compared to control. Post-generator ΔP was significantly lower than control only at hours 8, 12, and 17. The mean frequency at maximum power was similar for all circuit configurations (Figure 2 A – F).

**Figure 2.**
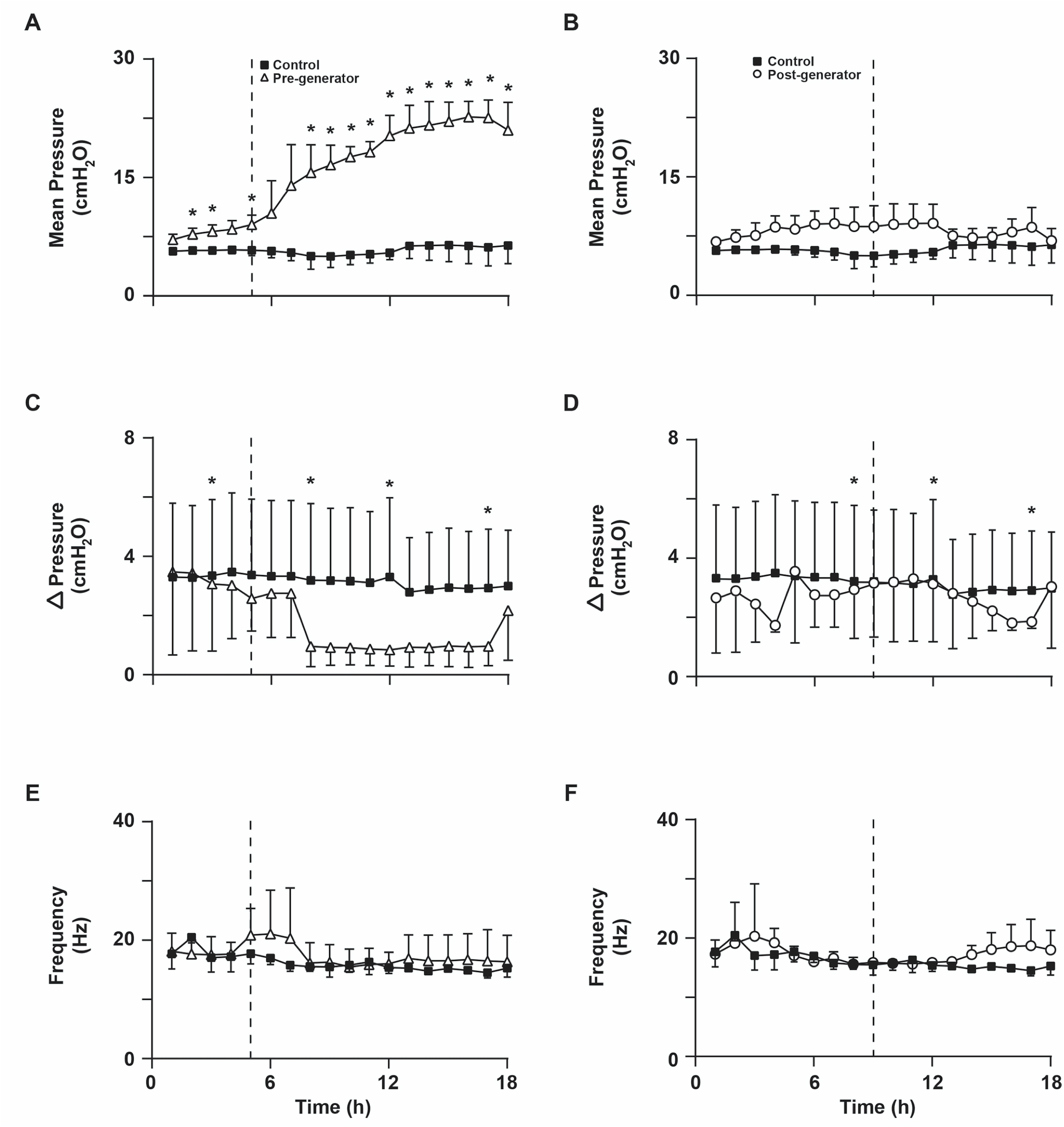
Mean pressure (A-B), ΔP (C-D), and frequency (E-F) over time for the filter positions and control: No filter (None, black square); Pre-generator (Pre, open triangle), post-generator (Post, open circle). Data points are mean (SD) for n=3/group. The fail-points for the respective filter position are represented by a vertical dashed line. * p<0.05.

### Filter weight

Filter weights before and after use are reported in Table 1. for both circuit filter positions. The filter weight was significantly heavier in the pre-generator compared to the post-generator position at the end of the study. Significantly more water accumulated in the generator overflow in the post-generator positions compared to the pre-generator position and the control position. Inclusion of a filter did not alter the volume of water that accumulated in the CPAP circuit.

**Table 1.** Filter weights and volumes of water removed from the circuit.

|  | Filter Position |  |  |
| --- | --- | --- | --- |
|  | Control<br>(n=3) | Pre-generator<br>(n=3) | Post-generator<br>(n=3) |
| **Baseline** |  |  |  |
| Filter dry weight (g) | - | $8.8 \pm 0.1$ | $8.9 \pm 0.1$ |
| **End Study** |  |  |  |
| Filter weight (g) | - | $11.9 \pm 0.2^{\dagger}$ | $10.8 \pm 0.1^{\dagger\S}$ |
| Water removed from circuit (mL) | 4.0 (3.0) | 2.5 (3.5) | 2.0 (1.5) |
| Water removed from overflow (mL) | 0 (10) | 1 (6) | 160 (20) * |

Median (range); † p <0.05 compared to dry weight (baseline) for same filter position;
$\S$ p<0.05 compared to pre-generator at end study; \* p< 0.05 compared to control and pre-generator;

## Discussion

The placement of an electrostatic filter in a bubble CPAP circuit changes the stability of pressure delivery. Positioning of the filter either immediately before or after the bubble generator increases the mean pressure delivered above a predefined fail-point. Filter failure occurs earlier and is sustained when the filter is positioned immediately before the CPAP generator, compared to later and intermittent failure when placed in the post-generator position (point of exit of flow into the environment). Incorporation of a filter did not change the mean frequency of the maximum power in the oscillatory pressure waveform. The filter become more saturated with water when positioned in the pre-generator versus post-generator position.

Placement of a filter in a bubble CPAP circuit reduces water accumulation within the circuit; however, the filter increases in weight over time, indicative of water absorption by the filter. Filter saturation increases resistance [16]. This may explain the increased mean pressure and dampened pressure waveform with the filter in the pre-generator position. Microbial filters marginally increase the imposed work of breathing measured over 18 breaths across a range of non-humidified respiratory support devices [17]. In comparison, our recordings were in a humidified system and were conducted over an extended period (18 h); using this more clinically applicable setting we clearly show that time and humidification affect mean pressure when the circuit configuration includes an electrostatic filter, especially when the filter is positioned prior to the bubble generator.

A highly variable or noisy pressure waveform is characteristic of bubble CPAP. It contributes to the purported advantage for enhanced gas exchange due to the high-frequency oscillatory nature of the pressure waveform [7, 18]. The variable pressure is also fundamental for stochastic recruitment of collapsed alveoli [7-11]. Bubble CPAP improved measures of acute respiratory mechanics and reduced lung injury compared to constantly delivered CPAP in preterm lambs [7]. Damping of the pressure amplitude (ΔP) with the filter in the pre-generator position would reduce the efficacy of bubble CPAP for gas exchange and alveolar recruitment.

Although the *in-vitro* nature of this study may be considered a limitation, such models are used widely for describing the performance of respiratory support devices, including neonatal respiratory support modalities [15, 17, 19-22]. Our *in-vitro* experimental set-up was a sealed circuit, measuring the stability of pressure without considering leak. Clinically, bubble CPAP pressure delivery is determined by the bias flow and degree of leak at the nares, as well as the degree of mouth closure [15]. Many neonatal units aim to minimise leak by using chin straps (to keep the mouth closed) and colloid dressings over the nares that not only reduce nasal trauma but also markedly reduce leak at the nasal interface. The instability of the delivered pressure over time should concern clinicians considering the use of such filters when treating premature infants with bubble CPAP, particularly when positioning the filter immediately before the bubble CPAP generator chamber.

The transmission of SARS-CoV-2 to health-care workers is a major concern in the current pandemic [23]. Neonates are at risk of COVID-19, although evidence of infection is sparse. Newborns of mothers with confirmed or suspected COVID-19 require precautions as recommended in recent published international guidelines [24]. Filters reduced the expired viral load from ventilator circuits when tested in previous pandemics [25, 26] and are recommended by several international societies [27, 28]. However, it is essential to consider how such filters on impact the efficacy of the respiratory support and to identify required changes in clinical practice such as frequency of filter exchange, to ensure safety of both health-care workers and the patient.

We conclude that the addition of an electrostatic filter into the circuit of a bubble CPAP system must be undertaken with extreme caution. Placement of a viral filter in the expiratory circuit immediately before the bubble CPAP generator chamber is not recommended. The excessive drift in mean pressure delivery and damping of the pressure waveform with the filter in this position presents a safety risk for the infant that outweighs the benefit of preventing cross-infection. The addition of the filter at the flow exit point for the CPAP generator (post-generator) may be acceptable but requires monitoring of the pressure in the circuit at regular intervals. The manufacturer of this filter recommends changing it every 24 h. Based on the data presented, changing the filter at least every 12 hours, and likely every eight hours should be considered when used in bubble CPAP.

## Data Availability

Data is available upon request

## Statements

### Statement of Ethics

This paper is exempt from ethical committee approval as it was conducted in a laboratory without the participation of human or animal subjects.

### Author contributions

MJ Dahl and JW Davis made substantial contributions to the study conception and design, acquisition, analysis, interpretation of data, drafting and revising the manuscript.

MN Cooper made a substantial contribution to the analysis, interpretation of the data and drafting and revising the article for submission.

JJ Pillow contributed to the study design, analysis and interpretation of data, and made a substantial contribution in revising the manuscript and approving for submission.

